# An assessment of the Health Information System in Khomas region, Namibia

**DOI:** 10.1101/2024.06.28.24309648

**Authors:** N J V Jatileni, E Nicol

**Affiliations:** Division of Health Systems and Public Health, Stellenbosch University, Cape Town, South Africa; Directorate of Special Programs, HIV and TB, Ministry of Health and Social Services, Windhoek Namibia; Burden of Disease Research Unit, South African Medical Research Council, Cape Town, South Africa

**Keywords:** RHIS, NHIS, competencies, Namibia, Behavioural factors, PRISM framework, Data use

## Abstract

**Introduction:** A robust and well-functioning Health Information System (HIS) is crucial for managing patient care, monitoring health system performance, and informing public health decisions. However, Namibia, like many developing countries, faces challenges in its HIS, such as limited financial and human resources, knowledge gaps, inadequate infrastructure, and behavioural barriers such as resistance to adopting new systems and a lack of supportive policies. Previous studies have not shown significant improvements since 2012. This study in Namibia’s Khomas region aims to assess human factors affecting the HIS and evaluate progress made from 2012 to 2023. It will use recommendations from a 2012 assessment by USAID to provide insights and propose ways to enhance healthcare delivery and resource allocation.

**Methods and analysis:** This study utilizes a cross-sectional design employing a multi-method approach to evaluate the performance of the Health Information System (HIS). Qualitative methods include conducting 17 in-depth interviews with key informants, a retrospective document review from the Ministry of Health and Social Services headquarters in Windhoek, supplemented by a modified office/facility checklist from all 14 health facilities in the Khomas region. The quantitative methods involve administering a questionnaire to 330 staff members, utilizing an adapted version of the Performance of Routine Information System Management (PRISM)’s Organizational and Behavioural Assessment Tool (OBAT). Descriptive statistics will be applied to analyse the quantitative data, while a deductive interpretive approach will be used for qualitative data analysis.

**Ethics and dissemination:** The protocol was approved by the Stellenbosch University Health Research Ethics Committee (Reference No: S23/05/119), the Namibia ministry of Health and Social Services (Reference No: 22/3/2/1) and will adhere to the principles of the Declaration of Helsinki (1964). The study aims to identify barriers and facilitators for implementing recommendations across different levels of the Health Information System (HIS), with a focus on improving the HIS in the Khomas region. Outputs will include communicating the findings to the study population, presenting at both local and international conferences, and publishing peer-reviewed journal articles.

## Introduction

Health information is an important building block of the health system and is vital for the effective functioning of the other five building blocks - service delivery, health workforce, medical products, vaccines and technologies, financing as well as leadership and governance ^1^. Hence data generated from the health information system (HIS) should be fit for purpose and of good quality i.e., accurate, reliable, integrated and analysed in a timely fashion ^1^. A functional HIS involve a continuous process of information gathering, processing, transmission, storage, and management within the health sector ^1.3^ and should provide reliable information sources for decision-making especially for resource allocation as well as policy and strategic plannings to ensure the delivery of good healthcare services ^3^. Studies show that the quality of a good HIS is affected by the data collection methods and tools, the availability and knowledgeability of the staff using and collecting the information, as well as how and when the information is analysed ^1-4^.

The World Health Organization (WHO) together with other agencies such as United States Agency for International Development (USAID) and the United States President’s Emergency Plan for AIDS Relief (PEPFAR) play a vital role in assisting developing countries to upgrade their HIS^5-7^. These aids come in the forms of scholarships and workshops for the workforce, funding of programmes, purchase of equipment that aid in data collection and analysis at different levels of the HIS, as well as employing qualified staff and providing recommendations to improve the HISs ^5-7^. Despite the assistance from these agencies, low- and middle-income countries (LMICs) still battle to implement the recommendations ^3,8^, which mostly noted were to clearly define staff roles and structures, especially for HIS, support various training for staff and give incentives to staff to retain them ^3,5,7-11^.

## Background

Namibia, an upper-middle-income country whose national HIS is led by the Ministry of Health and Social Services (MoHSS), faces challenges similar to other developing countries ^9,10^. These challenges include insufficient HIS staff, absence of HIS policies, poorly defined staff roles especially for the health care workers taking on multiple roles including data capturing or management, as well as the absence or insufficient equipment ^2,5,8,10-15,25^. The most noted challenges in Namibia, however, were around the human-related component of the HIS such as insufficient HIS experts and staff to develop the HIS, lack of clearly defined roles and structures, insufficient staff training and untrained staff in scenarios with high staff turnover ^3,10-11,13,15^, in addition to staff resisting change ^15^. Other challenges highlighted were unintegrated data systems, where donors and the Ministry of Health have different data systems; unmonitored and un-evaluated systems implemented, lack of resources to support and sustain the implemented systems, political challenges, and absence of policies; as well as inadequate infrastructures such as computers and internet ^3,10-11,13,15^.

In 2012, USAID assessed Namibia’s national Health Information Systems (NHIS) to integrate and strengthen the system. The assessment aimed to take inventory of existing systems, identify strengths and weaknesses, and make recommendations for planning ^11^. Findings were categorized into four thematic areas: data and information; technology, protocols, and the human interface; information products, data use, and knowledge management; and management, coordination, and implementation. Challenges highlighted included resistance to new systems, inadequate change management strategies, lack of standardized data collection tools, insufficient managerial skills, poor work ethics, and shortages of human and technical resources at both local and Ministry of Health and Social Services levels ^11^.

The study noted that most challenges were human-related, particularly focusing on staff motivation and attitudes ^4,16^. It recommended forming a committee within the HIS directorate, tasked with strategic planning, stakeholder coordination, and leading the implementation of recommendations ^11^. Subsequent studies reported similar challenges, without clear evidence of improvement ^10,13^. An exploratory study conducted six years after the initial assessment in 2012 not only revealed a fragmented system but also identified significant human factor deficits ^2^. These included staff shortages, insufficiently trained personnel, and structural or organizational challenges ^2^.

The main recommendation from a 2012 USAID study regarding the human aspect of the Health Information System (HIS) was to recruit and train staff to strengthen HIS capacity ^11^. Other recommendations emphasized the necessity of various training programmes, including on-the-job, refresher, and periodic training, both short- and long-term ^11^. However, over a decade later, there is little evidence indicating whether these recommendations were implemented or if there was any change in behaviour within the Namibian Ministry of Health and Social Services (MoHSS). Recent studies on the Namibian HIS suggest that most challenges stem from human-related factors ^2,9-11,14^.

Many studies on HIS conducted in Namibia have similar recommendations regarding technical, behavioural, and organizational aspects of the HIS. However, these studies often lack clear demonstrations of system improvement over time, beyond the continual introduction of new challenges and information ^2,10-11,13,15^. Despite recommendations aimed at enhancing the quality of data produced by the National Health Information System (NHIS), challenges persist in the performance of the Regional Health Information System (RHIS), many of which are attributed to human factors. The extent to which the 2012 recommendations from USAID have been implemented and their impact on RHIS improvement remain unclear. Additionally, most evaluations of the Namibian HIS have not thoroughly investigated the human factors influencing it ^2,10-11,13,15^.

The proposed study aims to address gaps in HIS by utilizing the Performance of Routine Information System Management (PRISM)’s Organizational and Behavioural Assessment Tools (OBAT) alongside qualitative methods. It will assess behavioural improvements in the RHIS in Namibia’s Khomas region over the past decade and identify barriers and facilitators impacting the implementation of recommendations for enhancing the Namibian HIS. Additionally, the study aims to evaluate human factors influencing HIS performance in the Khomas region and assess overall improvement progress between 2012 and 2022, utilizing recommendations from the National Health Information Systems Assessment conducted by USAID in 2012.

These will be achieved through the following objectives:

1. To assess trends in HIS-related human resources allocations in terms of staff capacity, skills, and competencies in the Khomas region since 2012.
2. To identify the behavioural and organisational factors influencing the performance of the NHIS in the Khomas region.
3. To determine the barriers and facilitators of implementing the recommendations from the USAID assessment of the NHIS in the Khomas region.

## Materials and Method

### Conceptual framework

The PRISM framework is a robust tool for assessing and improving routine health information systems (RHIS) performance ^17^. It examines three key areas: behavioural, organizational, and technical components of RHIS. Behavioural determinants focus on staff motivation, including knowledge, skills, attitudes, and motivation. Organizational determinants address factors like resources, information culture, and management support. Technical determinants involve aspects such as data collection tools, processes, and systems availability ^17-18,19-23^. The framework comprises four tools designed to assess or evaluate RHIS. These tools include the Performance Diagnostic Tool, targeting technical determinants, and a Facility or Office Checklist, which assesses the link between resource availability and RHIS implementation at facilities, focusing on technical aspects. Additionally, the Management Assessment Tool (MAT) examines RHIS management practices to enhance overall HIS management. Finally, the Organizational and Behavioural Assessment Tool (OBAT) investigates behavioural and organizational factors influencing RHIS performance ^17-21,28^.

The PRISM framework and its tools have proven effective in enhancing HIS in numerous African countries, including Ethiopia ^24^, Uganda ^21^, and South Africa ^18^. However, there’s no evidence of their use in Namibia. Applying the OBAT tool to study behavioural and organizational determinants in the Khomas region would be particularly intriguing.

### Study design

The study design is a cross-sectional study employing a multi-method approach to assess the performance of the HIS in the Khomas region of Namibia. The multi-method approach includes both qualitative and quantitative methods. The PRISM’s OBAT questionnaire and Office/Facility checklist will be adapted and customized for the Namibian context, as it will be sufficient to address the behavioural and organizational factors promoting the use of health information ^8^. An adopted interview schedule from a study conducted in Senegal ^25^ will be used to conduct interviews with key informants. This interview schedule has two parts, one for the national level mid-level analyst and M&E personnel, and the second for high level decision-makers ^25^. Key informants will include high level decision makers, as well as mid-level analysts. Furthermore, a retrospective document review, guided by a checklist will be conducted. Documents to be reviewed will include the HIS policy, latest minutes of the HIS committee meetings, latest information on availability of support structures such as study leave, loans, or bursaries from MoHSS to improve staff knowledge and capacity.

### Study setting

This study will be conducted in the Khomas region, one of Namibia’s 14 regions. This region encompasses the Windhoek district, which serves as the capital city of Namibia (Figure 1). As per the 2018 situation analysis on human resources for health in Namibia, the Khomas region spans a geographical area of 36,964 km^2^, with an estimated population of 447,636. Additionally, it has a population to health facility ratio of 37,303 ^26^.

**Figure 1:**
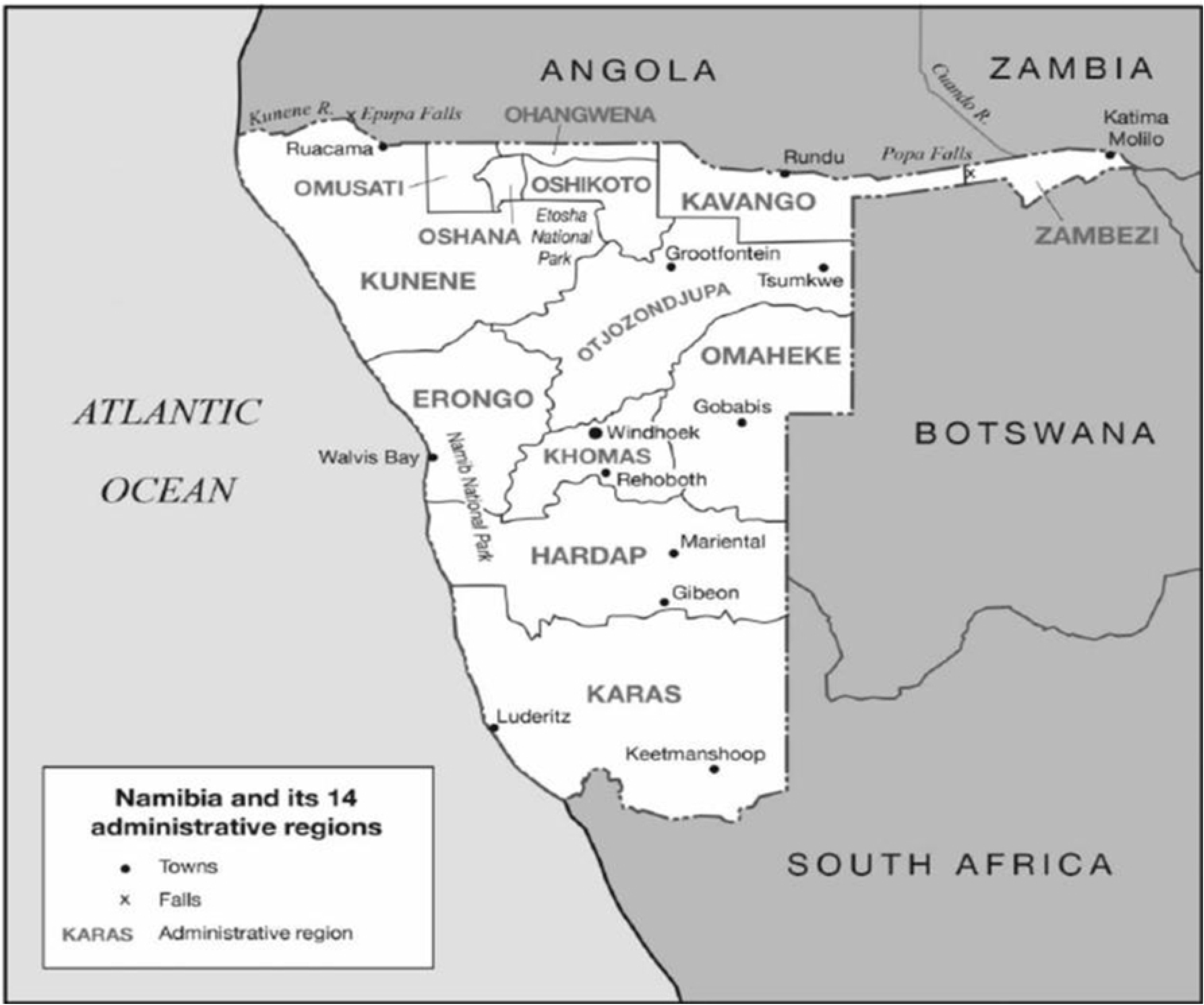
Khomas region, Windhoek district map. Source:^27^

**Figure 2:**
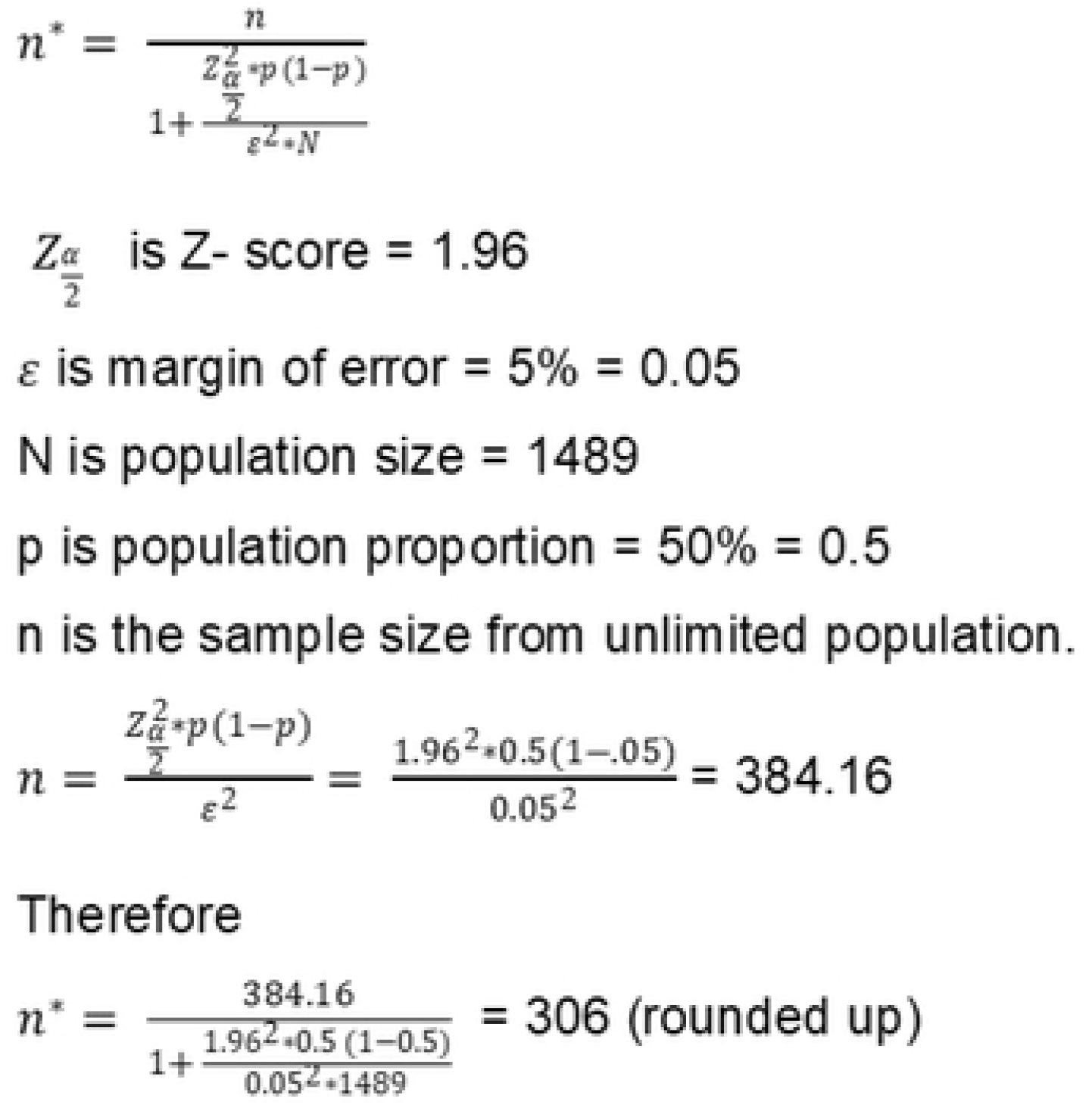
Sample size calculation from a finite population

### Health facilities

The Khomas region comprises 14 public health facilities, with 11 located in Windhoek and three on the outskirts within 40-110 km. These facilities include one national referral hospital (Windhoek Central Hospital), one intermediate hospital (Intermediate Katutura State Hospital), three health centers (Okuryangava, Khomadal, and Katutura Health Centers), and 10 clinics (Wanaheda, Robert Mugabe, Otjomuise, Maxuilili, Hakahana, Donkerhoek, Dordabis, Groot Aub, and Baumgartsbrunn Clinic). This region was selected due to its high patient to staff ratio, diverse health facility types, and the presence of the national Health Information and Research Directorate of the Ministry of Health and Social Services, facilitating easy access to information.

### Data collection and analysis

Data will be collected from all the 14 health facilities in the Khomas region as well as from the health information systems department at the Ministry of Health and Social Services headquarters in Windhoek electronically using SUN survey. Data will be collected by the principal investigator and two research assistants, who will be extensively trained on the OBAT questionnaire, the office/facility checklist, the retrospective document review checklist, and interview schedules. The training will also include interviewing skills, proper recording of answers to ensure accurate and consistent data.

### Study design, data collection and analysis

#### Objective 1

We plan to assess trends in HIS-related human resources allocations in terms of staff capacity, skills, and competencies in all 14 health facilities in the Khomas region since 2012. This comparative and exploratory study will use both qualitative and quantitative methods:

a. Qualitative: Retrospective document review of HIS policies, minutes from HIS committee meetings, and information on study leave, loans, and bursaries.
b. Quantitative: Facility survey of health information personnel and resources in the Khomas region health facilities and the Health Information and Research Directorate (HIRD) at the Ministry of Health and Social Services (MoHSS).

A retrospective document review will be conducted to assess trends in the availability of healthcare workers in selected facilities from 2012 to 2022. This review will include HIS policies, the latest minutes from HIS committee meetings, and information on study leave, loans, and bursaries. The data will be compared across health facilities and against staff establishments (the total number of approved jobs or posts in an organization) to determine overall improvements. Additional data to be assessed will include the availability of an HR information system (manual or electronic), training records or proof of training, policies, HIS committees and directorates, and support structures like study leave, loans, or bursaries provided by the MoHSS to enhance staff knowledge and capacity. These factors will be compared from 2012 to 2022 to identify any changes over time.

This document review will be conducted simultaneously with in-depth interviews with HIS department cadres, including the HR department. Information will be collected for different years to evaluate overall improvements in the HIS system. If older information is not accessible, the latest available data will be used. A modified PRISM office/facility checklist will be administered at all 14 health facilities and the HR department to examine HR cadre availability and training records over time, using the latest available data if older information is unavailable.

The analysis of the document review will be carried out through a deductive content analysis, which involves interpreting the information contained in the documents and drawing conclusions and recommendations. To facilitate this process, a document review checklist has been developed, which includes four categories that will be used to gather and analyse the required documents. These categories are listed in the document review checklist.

To ensure that the information collected is properly tracked and organized, Microsoft Excel will be used as a tool to manage the documents reviewed. The documents will be categorized based on their dates and any other relevant metadata. Additionally, detailed notes will be taken to highlight the current structure, functionality, and patterns observed over the years, as well as any other relevant information that will aid in drawing conclusions and recommendations.

Data from the Office/Facility checklist will be exported onto STATA Version 16 and analysed using the ANOVA two way. Dependent and independent variables will be assessed, and graphs such as bar charts or histograms will be generated to assess trends from 2012, in terms of staff turnover, training, experience or education, as well as show and compare the organizational and behavioural factors. Also, the average absolute deviation from the mean will be calculated for each year to identify consistency the data.

#### Objective 2

We will conduct a comparative and an analytical observational study to identify the behavioural and organisational factors influencing the performance of the NHIS in the Khomas region. A quantitative of data collection, which will include a self-administered questionnaire adapted from the PRISM’s Organizational and Behavioural Assessment Tool (OBAT) for the participants in the Khomas region health facilities and the Health Information and Research Directorate (HIRD), at the Ministry of Health and Social Services (MoHSS). The questionnaire will be used to assess participant’s behaviours, attitudes, and knowledge, and to determine the factors influencing the performance of the NHIS in the Khomas region.

The sample size calculation depicted in Figure 1 was conducted with the assistance of a statistician. This phase of the study’s sample size was determined based on the estimated total of 1513 healthcare workers in the Khomas region. This includes 1489 nurses, divided into 522 enrolled nurses (E/N) and 967 registered nurses (R/N), along with 24 data management staff, comprising 15 data clerks, five health information systems (HIS) officers, and four staff in the health information systems department, resulting in a total of 330 participants (i.e., 306 healthcare workers and 24 data management staff).

The 306 participants will be randomly recruited between 1 July and 30 September 2024 until the maximum sample size is reached. These sampling methods are designed to ensure minimal bias. These staff members are distributed across all 14 health facilities, outreach services, and are employed by either the Ministry of Health and Social Services (MoHSS) or funded by organizations such as PEPFAR, I-TECH (International Training and Education Centre for Health), and the Global Fund.

The PRISM’s OBAT questionnaire will be administered to 330 participants, including 306 nurses, 15 data clerks, five HIS officers, and four staff from the HIS department. It comprises four sections tailored to different levels of staff: staff and management at all levels, district and higher levels, health facility in-charge, and data management staff. These sections aim to identify organizational and behavioural factors influencing the HIS systems. The results will provide insights into overall improvement.

To ensure data reliability, the investigator/researcher will randomly check captured data. Captured data will be extracted into Microsoft Excel and compared using the Excel Comparison tool. Any inconsistencies will be identified and validated against the captured questionnaires. Further data cleaning and validation will involve generating frequency tables for each data element to identify missing or inconsistent values that may have occurred during data capture.

Data from the OBAT questionnaire will be exported to STATA Version 16 for analysis using the ANOVA two-way method. The analysis will involve assessing both dependent and independent variables. Graphs, such as bar charts or histograms, will be generated to visualize trends from 2012, focusing on factors such as staff turnover, training, experience, and education, as well as organizational and behavioural factors. Additionally, the average absolute deviation from the mean will be calculated for each year to identify data consistency.

#### Objective 3

This study utilizes an explanatory design, relying on qualitative methods of data collection. Specifically, it involves in-depth interviews with key informants to determine the barriers and facilitators of implementing the recommendations from the USAID assessment of the NHIS in the Khomas region.

Key informants for this study will include personnel from all fourteen health facilities in the Khomas region, as well as personnel from the Ministry of Health and Social Services Health Information Systems. Participants must be stationed at public Ministry of Health and Social Services (MoHSS) health facilities in the Khomas region. They can be either employed by MoHSS or funded by donors. To be eligible, respondents must have worked with HIS data at the health facility for a minimum of 6 months, including data entry staff, facility in-charges, and managers in the HIS departments. Staff employed at private health facilities will be excluded from the study.

A purposive sampling method will be employed to select 17 participants for the interviews. This sample will consist of a senior nurse or in-charge nurse participant from each of the 14 health facilities, one senior or team lead data clerk, another senior health information systems officer, and one cadre from the HIS department.

In-depth interviews will be conducted face-to-face or virtually between August and November 2024 with 17 key informants from health facilities and the Health Information and Research Directorate using an interview schedule. The interview schedule comprises two parts: the first part contains questions for the national level HIS department, administered to one person at this level. The second part includes questions for mid-level analysts and M&E personnel, administered to the remaining 16 participants, comprising 14 in-charge or senior nurses at each health facility and two district and regional health information officers. The interviews, scheduled to last between 20-30 minutes, will be voice-recorded and conducted in both English and the local languages (Oshindonga and Afrikaans) spoken at each site. These recordings will be translated to English before analysis. Observational notes on participants will be taken when necessary and used to complement the in-depth interviews.

The voice-recorded data will be transcribed and coded using ATLAS.ti. Transcriptions of the Oshindonga and Afrikaans interviews will be translated into English. The principal investigator and assistant researchers will carefully review the transcripts to ensure accuracy and completeness, with special attention to capturing all relevant codes. A deductive thematic approach will then be employed to analyze the themes and identify barriers and facilitators related to implementing recommendations from the USAID assessment of the NHIS in the Khomas region. Table 1 outlines the connections between the study objectives and the data collection tools.

**Table 1:** Linkages between the study objectives and data collection methods.

| Data collection | **Objective 1**<br>(Trends assessment in HIS-related human resources) | **Objective 2**<br>(Identify behavioural and organisational factors) | **Objective 3**<br>(Barriers and facilitators) |
| --- | --- | --- | --- |
| **Quantitative methods** |  |  |  |
| Self-administered questionnaires with national and facility level staff (PRISM OBAT) |  | X |  |
| A modified/office or facility checklist | X |  |  |
| **Qualitative methods** |  |  |  |
| A retrospective document review (data review on staff trends from the system) | X |  |  |
| In-depth interviews with key informants |  |  | X |

### Pilot study

A pilot study will be conducted in another region, specifically at Okahandja District Hospital and Nau-Aib Clinic, where the same questionnaire will be administered to similar participants from hospitals and clinics. The pilot will include 10% of the total sample size, totalling 34 participants, consisting of thirty-one nurses, one district Health Information Systems (HIS) officer, and two data clerks. The primary aim of the pilot study is to assess the feasibility of the data collection tools for use in the Khomas region study. It ensures the clarity of questions, monitors completion time, and allows for corrections to be made before fieldwork, reducing bias, and ensuring a standardized collection tool. Any necessary amendments will be implemented before the full study. The pilot study guarantees that methods are suitable and will yield quality, reliable results.

### Ethical considerations

The study obtained ethical clearance from the Stellenbosch University Health Research Ethics Committee (Reference No: S23/05/119) and received permission from the Namibian Ministry of Health and Social Services (MoHSS) (Reference No: 22/3/2/1). Both verbal and written informed consent will be obtained from participants of the OBAT survey and in-depth interviews. The consent for in-depth interviews includes permission for recording. This ensures participants are aware of their rights, confidentiality measures, and the benefits of participation.

Data management will adhere to the Protection of Personal Information Act (POPIA). Data will be stored on a password-protected computer accessible only to the principal investigator. Voice recordings will be destroyed after research completion. Respondents and health facilities will be anonymized using unique codes (e.g., HF1 for health facilities, N1 for nurses). These facilities are suitable for confidential interviews, as they are the same ones used for patient management.

## Discussions

### Data dissemination

The results of this study will be disseminated to the Ministry of Health and Social Services, including the staff who participated in the research. The principal investigator will be available to present the findings if requested by the Ministry or other stakeholders. Furthermore, the study will be submitted for publication in a peer-reviewed journal, and the authors will be accessible to provide explanations of the findings to interested parties.

The inclusion of the PRISM’s OBAT tool, retrospective document review, and in-depth interviews in the study design will enhance its methodological rigor. This study is the first in the country to apply two PRISM tools, the OBAT and modified office or facility tools, to assess and evaluate the HIS system after a decade, offering valuable insights into progress and informing the region on the human aspects of the HIS. Additionally, it will shed some insight into the barriers and facilitators of implementing recommendations at various HIS levels.

However, the findings’ generalizability may be confined to the region due to its diverse healthcare workforce and better resource accessibility, particularly with its sole reference hospital. Moreover, the study’s focus solely on the human aspect and behavioural patterns of the HIS may overlook other important aspects, such as the availability of data collection tools.

## Data Availability

No datasets were generated or analysed during the current study. All relevant data from this study will be made available upon study completion.

## Acknowledgments

Not applicable

